# Immunization Staff Training Needs Assessment: An Overview Of Findings From Cameroon

**DOI:** 10.1101/2023.09.19.23295815

**Authors:** Amani Adidja, Sangwe Clovis Nchinjoh, Jessica Gu, Fabrice Zobel Lekeumo Cheuyem, Nadege Nnang Edwige, Marius Vouking, Jude Nkwain Muteh, Valirie Ndip Agbor, Mbanga Clarence Mvalo, Nsah Bernard, Budzi Michael Ngenge, Shalom Tchokfe Ndoula, Andreas Ateke Njoh, Pietro Di Mattei, Calvin Tonga, Yauba Saidu

**Author notes:** **Corresponding author** (SCN).

## Abstract

**Background:** In recent years, Cameroon’s Expanded Program on Immunization (EPI) has witnessed a sizable decline in its performance. Many stakeholders have cited weaknesses in human resources as one of the major drivers for the observed trend. In a bid to better understand and redress this situation, the EPI and its partners conducted a Training Needs Assessment (TNA) among central, regional, and district staff. The assessment aimed to quantify and characterize core capability gaps and leverage the findings to design an evidence-based capacity-building plan.

**Methods:** A descriptive cross-sectional study was conducted using aggregated data from a survey carried out among EPI staff from May to September 2016 across Cameroon’s health pyramid - central, regional, and district levels; and analysis was done using Microsoft Office Excel 2016.

**Results:** Over half of EPI staff had worked for less than three years in their current post, and roughly three-third of them did not receive pre-service training on vaccination. Additionally, about half of them had not received any form of in-service training on immunization. Supportive supervision was the most frequently cited topic for training, with a surprisingly higher need at the central level (80%). Financial incentive was not a primary motivating factor for learning. Approximately half of the respondents at all levels were not aware of onboarding materials. Most of the respondents identified multiple meetings, high staff turnover, and competing priorities as major barriers to learning. Only about half of surveyed staff reported participating in performance reviews, with nearly half of these reviews conducted when an opportunity arose.

**Conclusion:** There are still many gaps challenging the EPI goal achievement in Cameroon. Sustainably addressing these issues will require a comprehensive framing of capability-building activities. High-quality EPI human resources will boost the country’s vaccination performance and contribute to the reduction of infant mortality in the country.

## Background

Globally, immunization has been a remarkable tool for reducing morbidity and mortality among children since the creation of the Expanded Program on Immunization (EPI) in 1974 [1]. Many countries including Cameroon, a country in Central Africa, over the years have made significant progress in achieving critical vaccination targets. Indeed, Cameroon reported a significant rise in vaccination coverage for the 1^st^ and 3^rd^ doses of the Diptheria-Tetanus-Pertussis containing vaccines (DTP) from 54% and 62% in 2000 to 88.6 % and 95% in 2013, respectively[2]. This steady increase of over 30 points has been arguably due to consistent political will and commitment towards infant health, a high social acceptance rate, financial and technical support from development partners, and the implementation of the Reach Every District (RED) strategy [3], amongst others.

Despite this promising rise in vaccination coverage, in recent years, vaccination coverage in Cameroon has stalled with occasional progressive decline in performance down to 76% and 69% for DTP-1 and DTP-3, respectively, in 2021[2]. Among other things, anecdotal evidence suggests that this may have been a direct effect of new challenges faced by the program. This includes the need to expand and sustain coverages, close equity gaps dramatically, meet the demands of a rapidly rising birth cohort and urbanization[4], introduce new vaccines [5], and aggressively pursue accelerated global and regional disease control targets[6, 7].

To effectively deal with the immunization challenges of the 21st century, the EPI and its partners conducted several vertical vaccination system assessments and leveraged the findings to implement tailored interventions to close identified gaps. Although the implementation of these actions yielded positive results, the outcome is arguably limited by capacity gaps among EPI staff. Indeed, data from the 2016 national cold chain inventory showed that only 12.2% of the 9,661 surveyed staff had received training on immunization within the previous five years, a finding which is somewhat consistent with the 16% reported in 2013 [8]. This situation, coupled with a lack of updated reference documents, high staff attrition, and the absence of a robust onboarding system, have created significant capacity gaps among EPI staff at all health pyramid levels. As a result, many EPI staff, including logisticians, storekeepers, data managers, and vaccinators, lack the required skills to effectively do their job.

Although the EPI has invested significant efforts to address these challenges, five system issues have hindered progress toward building a competent workforce. First, the current training methods (notably, off-site classroom training) are mostly based on theoretical/didactical materials that are not always adapted to the Cameroonian context and that are delivered by instructors who are not sufficiently trained on the content. In addition, there are often no post-training follow-ups to ensure that training translates into improved performance. Secondly, there is often poor coordination of training between partners and the government, leading to poor training quality control and sometimes duplication of efforts. Third, partners often fund training activities, implying that capability-building activities can stall if partner funding evaporates. Fourth, the current training system is not viewed as a continuum to the extent that there are no clear links from pre-service to in-service to on-the-job training. Lack of this perspective often results in one-off training, which is driven by available funding for training of interest (e.g., new vaccines or technology introductions). Finally, there is no monitoring and evaluation system to track who has received/has not received training as well as knowledge utilization.

There is a need to establish a comprehensive capability program that can sustainably address current and future system challenges, including the high numbers of untrained staff, high staff turnover, and fast-paced and evolving technology. The first step towards establishing such a program necessitated the conduct of a Training Needs Assessment (TNA) to quantify and characterize existing knowledge and skills gaps among EPI staff at all system tiers. In this paper, we present the findings of our study that assessed existing capability needs among EPI staff and proposed strategies that can be leveraged to address the identified gaps to inform actions or interventions from policymakers, funders, and other interested immunization stakeholders.

## Methods

### Study design and setting

A descriptive cross-sectional study was conducted using aggregated data from a survey carried out among EPI staff from May to September 2016 across Cameroon’s health pyramid - central, regional, and district levels to establish vaccination training needs.

### Study population and sampling

Our study was based on aggregated data unveiled by the survey conducted from May to September 2016 among EPI staff from central, regional, and district levels of Cameroon’s health pyramid. For the purpose of research, this data was accessed in March 7^th^, 2023 and authors did not have access to information that could identify individual participants. The aggregated data contained a summary outcome of interviews of staff at central and regional levels, excluding non-technical staff such as drivers, cleaners, and security personnel. At the district level, available data was on staff considered during the survey through purposive sampling to assure proportional distribution rather than obtaining high precision. The aggregated data represents the view 28 EPI staff at central level, 52 at regional and 162 at district-level from 68 out of 200 health districts in Cameroon.

### Inclusion and Exclusion criteria

All data from the need assessment survey conducted in 2016 representing the view of technical staff working in the immunization space at the three target levels (central, regional, and districts) was considered. At the national level, this included staff from all existing functional departments, including the EPI manager and deputy, and staff from the following departments: administration and finance, routine immunization and logistics, data management, monitoring and evaluation, and communication for development. At the regional level, it included the regional EPI leads, logisticians, data managers, and surveillance and communication officers. At the district level, the district medical officers, the chief of the health bureau, and EPI focal staff where included in the final analysis. At all levels, we excluded support staff such as drivers, security guards, cleaners, and those working at the secretariats without a defined role as well as technical staff who did not participated in the survey.

### Data collection and procedure

A structured excel based tool was used to extract information from aggregated data on the need assessment survey. The final tool consisted of eight sections, including basic demographic information, vaccine-specific training, core skills and competencies, the preferred format of learning, motivational factors for learning, managers’ views on the current skill levels of staff under their command, human resource onboarding system at each level and skill/knowledge assessment on key immunization program concepts and practices.

### Data processing and analysis

Once the extraction was completed, it was assess for accuracy and completeness and cleaned. Data were then exported, processed, and analyzed using Microsoft Office Excel version 2016. Results were presented using frequency and percentages for the various variables of interest.

## Results

Overall, the aggregated data represents the view of 28 EPI staff at central level, 52 at regional and 162 at district-level from 68 out of 200 health districts in Cameroon. The majority of people working in EPI had less than three years of experience in their current posts, as shown in Table 1. Moreso, our findings showed that approximately half of the respondents at all levels (central: 55%; regional: 46%, and district: 49%) were not aware of onboarding materials for a new staff. In addition, most respondents had not received any form of pre-service training, notably 79 % at the central, 72 % at the regional, and 60 % at the district levels. Also, about half of the respondents at the district (50%), region (37%) and central levels (58%) had not received in-service training (Table 2).

**Table 1:** Staff professional experience across the three administrative levels of the EPI in Cameroon, 2016.

| Professional experience* | EPI level |  |  | Total |
| --- | --- | --- | --- | --- |
|  | n (%) |  |  |  |
|  | District<br>n=162 | Regional<br>n=52 | Central<br>n=28 | n=242 |
| **In all sectors** |  |  |  |  |
| < 3 years | 10 (6) | 2 (4) | 2 (8) | 14 (6) |
| 3-6 years | 36 (22) | 5 (10) | 16 (58) | 57 (24) |
| >6 years | 116 (72) | 45 (76) | 10 (34) | 171 (70) |
| **In immunization services** |  |  |  |  |
| < 3 years | 21 (13) | 7 (14) | 15 (54) | 42 (18) |
| 3-6 years | 45 (28) | 16 (30) | 9 (31) | 70 (29) |
| >6 years | 96 (59) | 29 (56) | 4 (15) | 129 (53) |

*\*Professional experience was defined as the total number of years an individual has worked following graduation from a training institution/university excluding internships.*

**Table 2:** Distribution of EPI staff according to the type of training received on immunization, 2016.

| Type of training received on immunization | EPI level |  |  | Total |
| --- | --- | --- | --- | --- |
|  | *n* (%) |  |  |  |
|  | District | Regional | Central |  |
|  | *n*=162 | *n*=52 | *n*=28 | *n*=242 |
| **Pre-service** |  |  |  |  |
| No | 97 (60) | 37 (72) | 22 (79) | 156 (64) |
| Yes | 52 (32) | 12 (24) | 5 (17) | 69 (29) |
| Can't remember | 13 (8) | 3 (4) | 1 (4) | 17 (7) |
| **In-service** |  |  |  |  |
| No | 81 (50) | 19 (37) | 15 (58) | 115 (48) |
| Yes | 73 (45) | 32 (61) | 13 (42) | 118 (49) |
| Can't remember | 8 (5) | 1 (2) | 0 | 9 (3) |

As shown in Table 3, cold chain logistics was the most covered topic during in-service training of EPI staff - 90 % of district, 83 % of regional, and 54 % of central-level EPI staff received in-service training on this subject. Other important topics including but not limited to Adverse events following immunization (AEFI), surveillance of vaccine-preventable diseases, conducting coverage surveys or Supplementary Immunization Activities were less prioritized for training. On the other hand, cross-disciplinary skills necessary for day-to-day operations seem not to receive sufficient attention during in-service training at all levels; for instance, less than half of district and regional staff received training on planning, budgeting, and report writing.

**Table 3:** Distribution of EPI staff according to the type of topic covered during in-service training by level, 2016.

| Topic | EPI level |  |  | Total |
| --- | --- | --- | --- | --- |
|  | *n* (%) |  |  |  |
|  | District | Regional | Central | *n*=118 |

|  | *n*=73 | *n*=32 | *n*=13 |  |
| --- | --- | --- | --- | --- |
| **Core immunization topic** |  |  |  |  |
| Cold chain logistics | 66 (90) | 27 (83) | 7 (54) | 100 (85) |
| Adverse events following immunization | 57 (78) | 20 (63) | 0 | 77 (65) |
| Surveillance of vaccine preventable disease | 55 (75) | 20 (63) | 0 | 75 (64) |
| Vaccine immunology | 38 (52) | 17 (53) | 4 (31) | 59 (50) |
| Community engagement | 44 (60) | 22 (70) | 0 | 66 (56) |
| Conducting supplementary immunization activities | 32 (44) | 16 (50) | 0 | 48 (41) |
| Conducting EPI Survey | 38 (52) | 17 (53) | 4 (31) | 59 (50) |
| Supportive supervision | 54 (74) | 26 (80) | 1 (10) | 81 (68) |
| Monitoring and evaluation | 51 (70) | 19 (60) | 0 | 70 (59) |
| **Other competencies (soft skills)** |  |  |  |  |
| Planning (including microplanning) | 31 (43) | 31 (60) | 0 | 62 (52) |
| Preparing proposals and report writing | 23 (32) | 16 (30) | 0 | 39 (33) |
| Use of Excel, Word, or PowerPoint | 17 (23) | 26 (50) | 0 | 43 (36) |
| Data management, reporting or analysis | 38 (52) | 36 (70) | 0 | 74 (63) |
| Budgeting & financing | 21 (29) | 22 (43) | 0 | 43 (36) |

### Preferred areas of training: Core immunization topics and transferable skills

Respondents indicated the need for training in all thematic areas but with varying frequency across themes and levels. Supportive supervision was the most frequently sorted topic for training, with a higher need expressed at the central level (80%). District and regional staff considered adverse events following immunization (AEFI), maintenance of cold chain equipment, stock management and distribution, and community engagement as critical areas where training is needed. Moreso, Over 50% of staff at all levels expressed the need to be trained in preparing reports, creating work plans, delegating tasks, time management, data analysis, and preparing PowerPoint Presentations. Similarly, over half of the respondents at all levels expressed interest in gaining skills in core aspects of managing teams/working in teams. Furthermore, over 40% of respondents at all levels highlighted the need to learn how to account for project-related expenditures/expenses (Table 4).

**Table 4:** Preferred areas for training on cross-disciplinary skills among Cameroon EPI staff, 2016.

| Preferred transferable skill | EPI level |  |  | Total |
| --- | --- | --- | --- | --- |
|  | *n* (%) |  |  |  |
|  | District | Regional | Central |  |
|  | *n*=152 | *n*=45 | *n*=30 | *n*=227 |
| **Oral communication** |  |  |  |  |
| Communication in public | 87 (57) | 30 (66) | 20 (65) | 137 (60) |
| Negotiation and advocacy | 59 (39) | 11 (24) | 14 (46) | 84 (37) |
| Mentoring | 49 (32) | 10 (22) | 7 (23) | 66 (29) |
| Training/teaching skills | 68 (45) | 19 (43) | 11 (35) | 98 (43) |
| **Written communication** |  |  |  |  |
| Report writing | 100 (66) | 35 (78) | 20 (65) | 155 (68) |
| Academic/research writing | 35 (23) | 9 (21) | 13 (44) | 57 (25) |
| Proposal/grant writing | 24 (16) | 7 (15) | 11 (38) | 42 (19) |
| Email communication | 73 (48) | 26 (58) | 21 (69) | 120 (53) |
| Formal memos/Government letters | 50 (33) | 22 (49) | 20 (65) | 92 (41) |
| **Project management** |  |  |  |  |
| Creating budgets | 35 (23) | 17 (37) | 16 (54) | 68 (30) |
| Monitoring budgets | 38 (25) | 21 (46) | 14 (46) | 73 (32) |
| Creating workplans | 90 (59) | 27 (61) | 19 (62) | 136 (60) |
| Monitoring workplans | 84 (55) | 27 (60) | 16 (52) | 127 (56) |
| Creating monitoring and evaluation plans | 73 (48) | 13 (29) | 16 (52) | 102 (45) |
| Accounting for expenditures/expenses | 62 (41) | 20 (44) | 12 (40) | 94 (41) |
| **Managing and working with teams** |  |  |  |  |
| Delegating tasks to others (productive task shifting) | 76 (50) | 22 (50) | 19 (62) | 117 (52) |
| Handling team challenges and conflicts | 62 (41) | 16 (36) | 21 (69) | 99 (44) |
| Juggling multiple activities/time management | 90 (59) | 24 (54) | 17 (58) | 131 (58) |
| Providing feedback to team members | 88 (58) | 25 (56) | 21 (69) | 134 (59) |
| Identifying poor performers and supporting improvement | 82 (54) | 26 (58) | 13 (42) | 121 (53) |
| **Data analysis and use** |  |  |  |  |
| Analysing data, making graphs, charts and tables | 81 (53) | 24 (53) | 14 (46) | 119 (52) |
| Using Excel, Access and/or other databases | 53 (35) | 14 (31) | 11 (35) | 78 (34) |
| Interpreting data and making decisions | 99 (65) | 28 (63) | 16 (52) | 143 (63) |
| Turning data evidence into action plans | 71 (47) | 15 (33) | 16 (52) | 102 (45) |
| **Designing knowledge products** |  |  |  |  |
| Creating PowerPoint Presentations | 74 (49) | 26 (58) | 15 (50) | 115 (51) |
| Creating research/survey protocols & tools | 27 (18) | 6 (14) | 11 (35) | 44 (19) |
| Creating job aids and/or training materials | 49 (32) | 12 (27) | 1 (4) | 62 (27) |
| Creating advocacy materials | 29 (19) | 7 (16) | 11 (38) | 47 (21) |

### Key challenges and drivers of learning

Surveyed staff largely aligned on four key factors that could impact learning – multiple meetings, high staff attrition, competing priorities, and team conflict (Table 5). With regards to motivation, financial incentive to learn was not among the leading motivation factors for all levels, contrary to what was previously thought. Other factors, such as obtaining a certificate of completion, being recognized by the ministry, and consideration for promotion or attending an international meeting, received better ratings than financial incentives at all levels. Classroom learning at the workplace and On-the-job mentorship and direct supervision were the top preferred learning methods in this study.

**Table 5:**
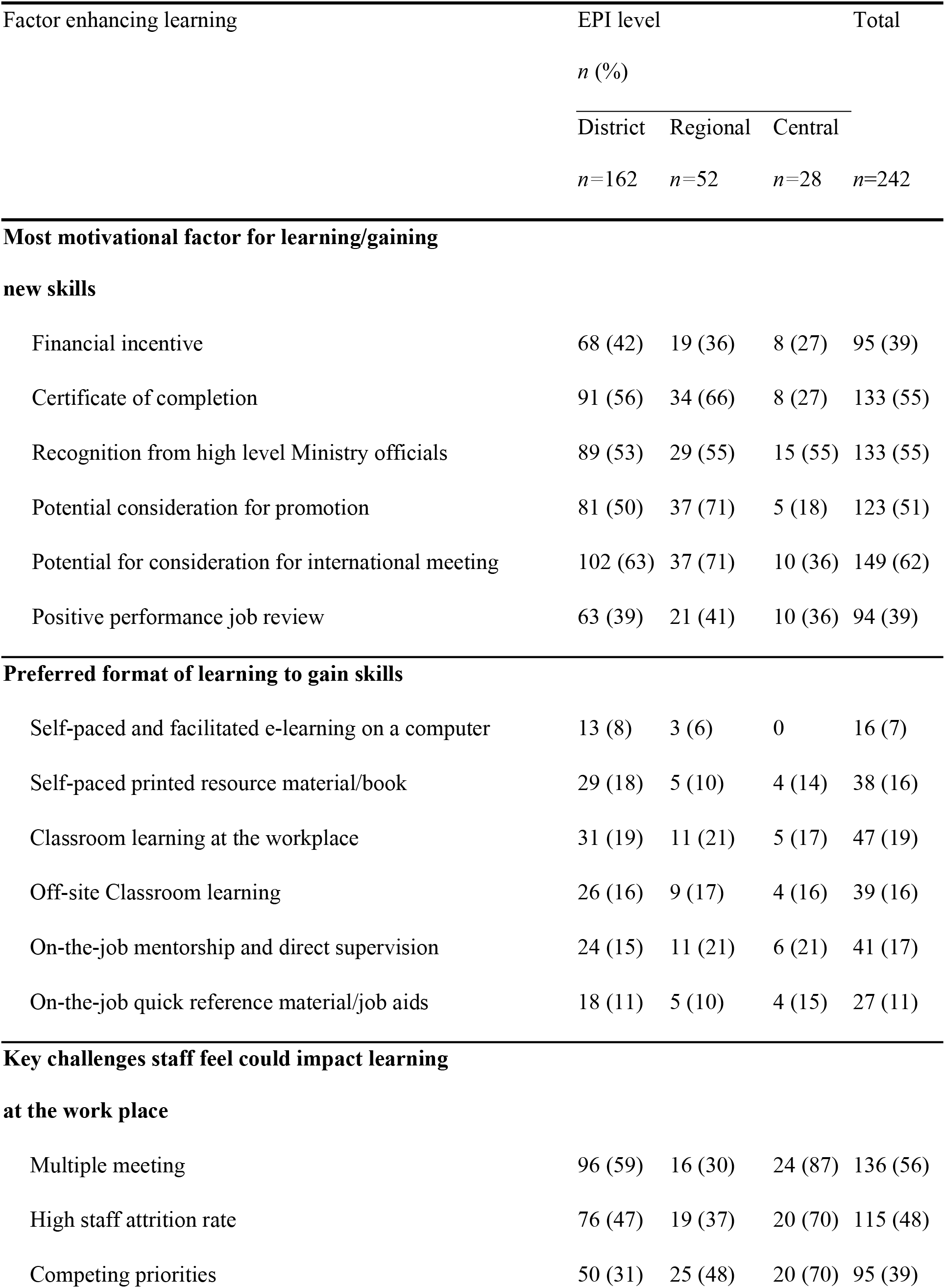
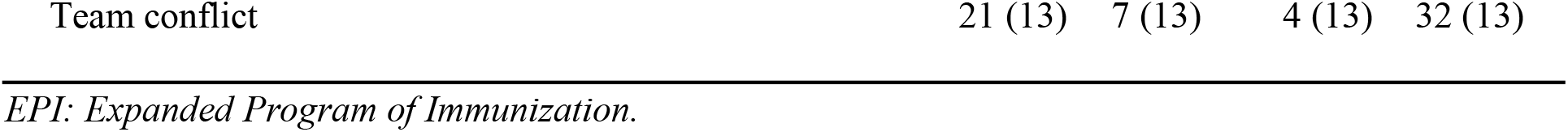
Self-reported factors that could enhance learning among Cameroon EPI staff, 2016.

## Discussion

In this study, the majority of people working in EPI had less than three years of experience in their current posts (Table 1), with approximately half of the respondents not aware of onboarding materials for new staff. This observation suggests high staff turnover, highlighting the need to strengthen the use of onboarding materials which is critical to sustainability following EPI-related capacity-building efforts. Robust onboarding can dramatically improve the performance and readiness of a person who takes on a new role in the health system. Onboarding helps build and sustain high-performing staff and maintains institutional knowledge. Therefore, onboarding materials are an essential tool of a well-functioning health system that can retain knowledge as staff changes[9]. The sub-optimal use of onboarding material despite high staff turnover arguably contributes significantly to the rising number of zero-dose and under-immunized children in Cameroon. This finding is consistent with a study conducted by Emily and colleagues in Tchad and Cameroon, which highlighted the role of rapid staff turnover in implementing Gavi’s health system strengthening support in both countries[10].

Moreso, the low vaccination capacity gap of EPI staff stemming from rapid staff turnover is further compounded by sub-optimal pre-service and in-service training at all health system levels (Table 2). This finding suggests that limited attention is being accorded to immunization in the curricula of nursing and medical schools (pre-service training). This is consistent with a study conducted at health training institutions in 12 target countries which suggested incomplete or outdated pre-service curricula, insufficient time allocation, and insufficient knowledge of the current EPI theory and practice by lecturers[11]. Redressing this situation will, therefore, require high-level advocacy and negotiations between the Ministries of Health and the ministries that govern professional education, such as the ministries of higher education and vocational training [12]. However, onboarding systems within the EPI need to be strengthened to provide newcomers with the necessary skills and competencies to effectively do their jobs. The finding that a sizable proportion of staff at all levels have received in-service training is reassuring in the sense that it indicates that mechanisms/systems are in place which can support in-service training. Pre-service training is fundamental in the skills acquisition of EPI staff; however, they require additional in-service training, onboarding, and supportive supervision to carry out their vaccination tasks efficiently[13].

Table 3 emphasized the knowledge gaps of EPI staff, with topics such as AEFI, surveillance, Supplementary Immunization Activities, and cross-disciplinary skills such as planning, budgeting, and report writing less frequently conducted. It is worth highlighting that the percentages reported for the central level need to be viewed with caution for two reasons. First, some degree of functional specialization is expected at the central level, and therefore, it will not be uncommon to see that certain staff have not received in-service training on some aspects of immunization. For instance, finance staff are not expected to receive in-service training on cold chain or conducting EPI coverage surveys because they may not require this knowledge for their job. Second, of the 28 central staff interviewed, only 13 indicated that they received in-service, making it difficult to draw any meaningful conclusion from the observed findings. Despite this limitation, staff at this level are expected to have a solid mastery of cross-disciplinary skills, given their role in evidence-based strategy development. And the fact that none of the respondents from the central level reported being trained on most topics suggests that in-service training are rarely tailored to provide staff with the needed competencies to do their jobs effectively. Therefore, there is a need to develop a more comprehensive and tailored capacity-building plan that meets EPI staff’s training needs sustainably, especially as respondents indicated the need for training in all thematic areas but with varying frequency across themes and levels (Table 4).

In this study, respondents considered self-paced printed resource material/book, classroom learning at the workplace, off-site classroom learning, on-the-job mentorship and direct supervision, and on-the-job quick reference material/job aids as top learning and work aids that could be employed to improve their skills. However, based on their perspective, self-paced and facilitated e-learning on a computer was the least effective. In all, the emphasis is on repetitive practical training approaches and regular supervision, consistent with evidence developed by Julia et al. on recommended in-service training techniques based on an integrative literature review[14].

Even with a state-of-the-art platform to support in-service training, some critical factors which pose serious hindrances to learning. Our study evaluated two core factors: staff motivation to learn and key challenges they feel could impact learning. With regards to motivation, financial incentive to learn was not among the leading motivation factors for all levels. Other factors, such as obtaining a certificate of completion, being recognized by the ministry, and consideration for promotion or attending an international meeting, received better ratings than financial incentives at all levels. In addition, over 30% of participants at all levels indicated that positive performance reviews from a manager or supervisor could motivate them to acquire new knowledge/skills. Overall, these findings suggest that strategies for training should also consider non-financial incentives, such as certification, recognition by senior officials, and the institution of a system that can guarantee regular performance job reviews. This will be important to promote (1) a culture of self-learning and (2) potentially work toward cost-savings of training methods (Table 5). However, surveyed staff aligned mainly on four key factors that could impact learning - multiple meetings, high staff attrition, competing priorities, and team conflict. While the impact of these factors on staff professional development could not be overemphasized, it is important to recognize that these factors could threaten the program’s overall performance. Therefore, addressing them-through optimal planning and prioritization-will enhance participants’ ability to learn and improve the program’s overall performance.

## Conclusion

Overall, this study suggests that most EPI staff at the surveyed levels lack the requisite disciplinary and cross-disciplinary skills to effectively do their jobs. These capacity gaps, to a large extent, stem from weaknesses in pre-service and in-service training systems. According to our findings, training needs vary considerably across the different levels and within the units/ department, and only limited time is available for professional development. This variation implies that efforts need to be invested in developing a tailored capability-building program, which covers relevant immunization topics and cross-disciplinary skills for each level, and which staff sees as useful for their jobs. Also, to effectively deal with the diversity of staff graduates from different training institutes at all levels, there is a need to institute a system that can guarantee targeted onboarding, mentoring, and performance review for new immunization staff. Our study raised crucial points on preferred learning methods and showed that factors such as obtaining a certificate of course completion are more motivating than providing financial incentives – which are useful for effectively planning EPI staff capability building programs.

## Recommendation

Based on our findings, we recommend immunization stakeholders:

1. Establish a competency framework that clearly defines the knowledge, skills, attributes, structures, roles and approaches needed for people within EPI to effectively do their job.
2. Leverage the competency framework to upgrade the capability of existing staff on core disciplinary and cross-disciplinary skills that are indispensable for their day-to-day operations.
3. Establish a robust onboarding system for new employees to ensure that updated technical and management/communication resources are available for each post/role.
4. Upgrade and standardize the current performance review mechanisms at all levels, as this can be a good opportunity to identify and address capability-building needs.
5. Strengthen pre-service training by ensuring that core immunization aspects are sufficiently integrated into the curriculum of, at least nursing and medical schools.
6. Enhance in-service training, whenever it is considered, by adapting and applying adult learning theories, strengthening the capacity of instructors, and appropriately selecting trainees, amongst others.
7. Establish a monitoring and evaluation framework to gather key information from every training event and track training effectiveness. Develop a routine database to gather this key information from every training event.

### Limitation

This assessment had four major methodological limitations. First, data were collected from 68 health districts, representing just 36% of health districts in the country. This limitation implies that this evaluation should not be considered a comprehensive presentation of the entire scope of capability gaps at the district level. Moreso, the study is based on a need assessment conducted in 2016; thus may not represent the current reality. Second, as per the objective of the TNA, this evaluation did not evaluate capacity gaps at the health facility level, where capacity needs mostly revolve largely around clinical practices. As such, the findings are not applicable to this level. Third, most of the findings of this evaluation are largely based on recall and perceptions. Fourth, some staff at the central level did not respond to some questions, which implies that the results for some thematic areas may not represent the variables explored. However, given the emphasis of the evaluation on pre-service and in-service training, with a lens on disciplinary and cross-disciplinary skills, the authors are confident that these limitations have not undermined the findings – the scope of data collection allowed the assessment team to draw out conclusions across nine thematic areas that may be applicable to central, regional and districts levels.

## Data Availability

Data is available upon administrative request.

## Author contributions

Conception & Design: AA, FZL, MV, JNM Statistical analysis: AA, JNM, MV, FZL Initial draft preparation: AA, SCN, JG, NNE Review and Editing: SCN, VNA, MCM, NB, BMN, STN, AAN, PDM, CT, YS Supervision: YS All Authors read and approved submission of the final article.

## Ethics approval and consent to participate

This study did not require ethical approval as it did not contain individual-level data collection.

## Availability of data and materials

Data used for this research is available from the corresponding author upon reasonable request.

